# PubChat: Evaluating the Effectiveness of Semantic Search in Establishing Drug Safety in Children^*^

**DOI:** 10.1101/2025.10.01.25335910

**Authors:** Ava Smith, Ella Dunbar, Camerron Crowder, Matthew Might

## Abstract

Children often respond differently to pharmaceuticals differently than adults, necessitating dedicated safety labeling. Although pediatric safety labeling is available through the FDA Online Label Repository, updates often lag newly published research, and manual searches of drug lists lack efficiency. To address these limitations, we developed PubChat, a tool that leverages semantic search and language models to extract, summarize, and categorize pediatric safety information from scientific literature, enabling rapid comparison with FDA-approved labeling. In a random sample of 80 drugs, PubChat agreed with FDA labeling in 41.25% of cases. In 23.75% of cases, it found supporting evidence that the FDA age recommendation was potentially too conservative. By transforming unstructured literature into structured, actionable insights, PubChat establishes a foundation for enhanced surveillance of pediatric drug safety and offers a complementary tool to bridge the gap between evolving research and static regulatory labeling.

## 1. Introduction

Children and adults represent distinct populations in medicine, a concept that extends back to the Hippocratic Corpus.^1^ However, this idea gained significant traction in pharmaceutical research and legislation in the late 20th century. Pharmacology research has demonstrated that how the body processes and responds to medications differs across age ranges and contributes to variability in drug responses. Additionally, these changes are not limited to the dichotomous distinction between “child” and “adult” but instead occur dynamically over the lifespan. These differences are driven by factors including, but not limited to, total body water composition, gastric pH, receptor sensitivity, and body weight.^2,3^ As such, it has become evident that a drug’s safety and efficacy profile should be investigated over multiple age ranges.

Historically, children have been underrepresented in clinical drug trials in the United States.^4,5^ Since the 1990s, several legislative milestones have culminated in a regulatory infrastructure for pediatric study requirements, drug labeling, and post-market safety reviews.^3^ Pediatric drug labels are currently available at the FDA Online Label Repository.^6^ However, several important challenges remain, from real-time availability of the most recent safety data to the absence of entries for off-label and investigational drugs.

Although the FDA updates the Online Label Repository with post-market safety information, updates are often delayed by years. These delays could result in some entries being incomplete. Additionally, the repository contains limited information about off-label drugs, which are drugs that are prescribed for unapproved indications, age groups, dosages, or administration routes.^7^ Off-label prescriptions are used more commonly in children than adults, leaving children at increased risk for adverse side effects.^8^ Limited information on off-label and investigational drug use raises additional concerns in vulnerable populations, including pediatric patients with rare diseases. For instance, 95% of rare diseases do not have an FDA-approved treatment, and off-label and investigative drugs are routinely prescribed in these cases.^9^ While prescription of off-labels and investigational drugs may be medically appropriate, prescribers face significant knowledge gaps that impede clinical decision-making.

To address these challenges, we have developed and tested PubChat, an artificial intelligence (AI)-assisted tool to compile and evaluate pediatric safety data from biomedical literature (**Figure 1**). PubChat combines semantic search to identify the most relevant abstracts, a large-language model (LLM) to generate structured summaries, and a hallucination-check that cross-references in-text citations to the article metadata. This approach provides prescribers with three advantages over the FDA database: safety and efficacy information for (1) off-label; (2) investigational; and (3) in general, more comprehensive and updated pediatric safety information for drugs. This automated approach allows providers to receive comprehensive safety information to make more informed recommendations for their clinicians, particularly in children.

**Figure 1.**
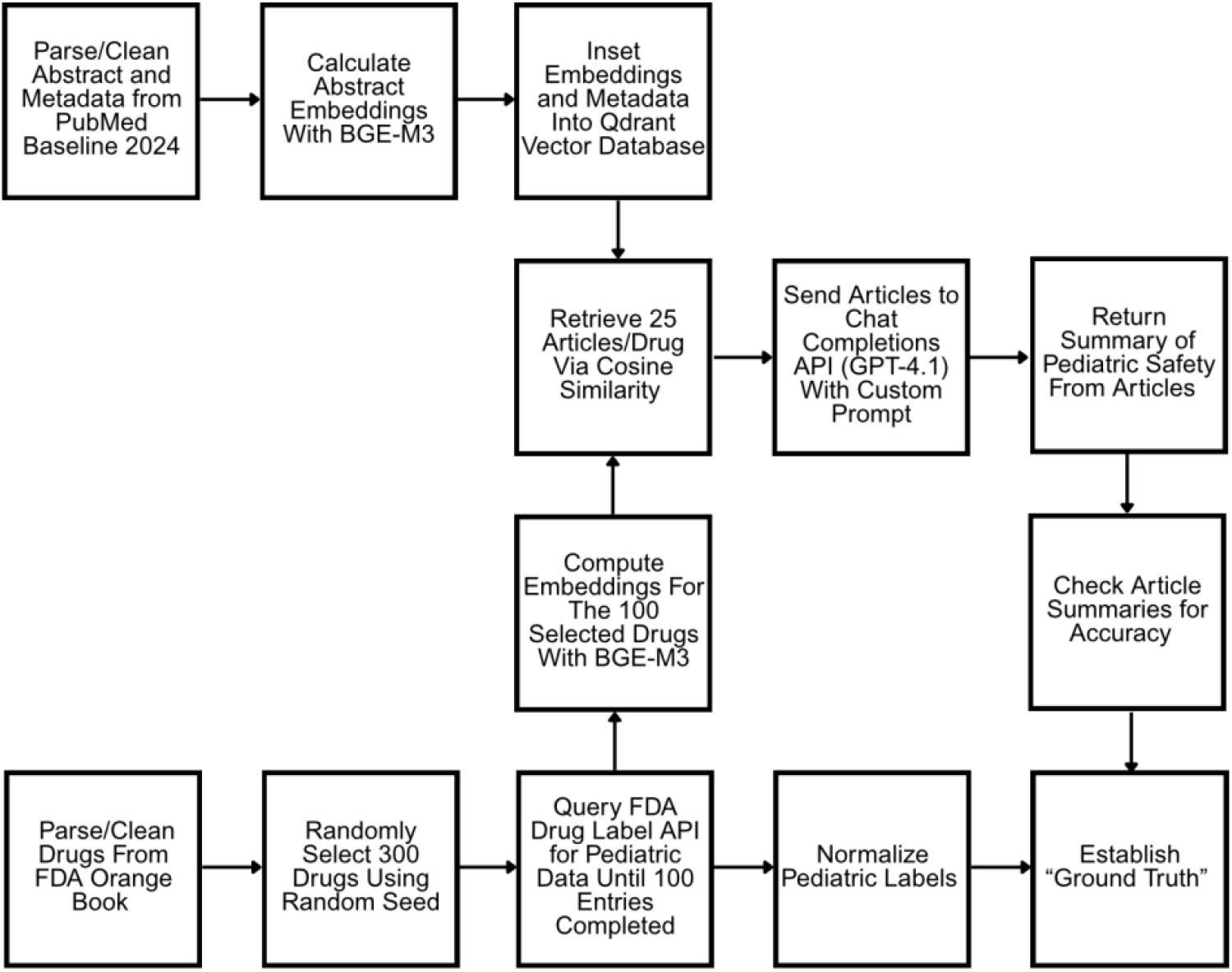
Summary of Pediatric Safety Evaluation Pipeline. PubMed baseline 2024 abstracts were parsed and cleaned, and their embeddings were computed using the BGE-M3 model and stored in a Qdrant vector database. A random sample of 300 drugs was drawn from which 100 were selected based on availability of pediatric data via the FDA Drug Label API. Embeddings for these drugs are also computed using BGE-M3 and normalized. For each drug, the top 25 semantically similar articles were retrieved using cosine similarity. These articles were analyzed using the GPT-4.1 Chat Completions API with a custom prompt to analyze pediatric safety information. The generated summaries were compared to official pediatric indications from FDA labeling to evaluate concordance with a 20/80 split between testing and training. Given the evidence for each drug, a “ground-truth” decision was made between the label in the FDA database, PubChat’s suggested label, or in cases where evidence did not support either label, neither. In-text citations generated by the GPT-4.1 were compared against article metadata to ensure citations were not hallucinated, and PubChat’s outputs were manually reviewed to ensure that articles were summarized appropriately.

## 2. Methods

### 2.1. Abstract and Vector Database Setup

The 2024 PubMed baseline repository, which contains all abstracts in PubMed, was accessed using the File Transfer Protocol (FTP).^10^ A custom Python script extracted abstracts and their associated metadata. Abstracts were selected over full-text publications because of their streamlined representations of published literature, ease of access, and scalability in a semantic search pipeline. Records with missing abstracts or that were marked as retracted were excluded. Each abstract was embedded locally using the BGE-M3 embedding model (dimensions = 1024) to create a semantic representation of the abstract.^11^

Next, each abstract embedding and its associated metadata were placed in a payload and uploaded to a Qdrant vector database.^12^ The database was configured with 1024 dimensions to match the embedding model and with “cosine similarity” as the comparison metric. The associated hardware was Mac Studio M2 with 192 GB RAM.

### 2.2. FDA Drug Label Dataset

Drug product data from the Orange Book, the FDA’s official publication of approved drugs, was obtained from the FDA’s website.^13^ Entries marked as discontinued and records with missing values in essential fields such as application number, application type, trade name, or active ingredient were removed. Each active ingredient had several entries corresponding to different manufacturers, routes of administration, and dosages. To construct a representative query set, each active ingredient was effectively given a weight of one, and 300 unique active ingredients were randomly sampled from the cleaned dataset using a reproducible random seed. All associated drug product entries for the sampled ingredients were then re-expanded, as the FDA Drug labeling API and Orange Book are not organized in one-to-one manner. The goal was to have one pediatric label per unique active ingredient, but sometimes multiple queries to the FDA Drug labeling API were necessary. Thus, the FDA Drug labeling API was queried for each active ingredient using the application number as a search term. The “trade name” in the Orange Book corresponded to the “brand name” in the Drug labeling API, and the “brand name” was extracted from the entry to ensure it matched the “trade name”. If the application number matched the name and the pediatric use section was not empty, the most recent label for the entry was saved and the program moved to the next ingredient. This was repeated until 100 total unique active ingredients were represented with a pediatric label. The first 20 entries were used as “training” data to develop the custom prompt, and the remaining 80 abstracts were used to test the model.

### 2.3. Abstract Retrieval and LLM Integration

For each drug, the term “safety of {drug} used in children” was embedded using BGE-M3, normalized, and compared to the stored vectors in Qdrant. 25 abstracts results per query were selected as a cutoff to allow for the most information to be supplied to the LLM while remaining under GPT-4.1’s context limit of one million tokens.^14^ Top-ranked abstracts were passed to the Chat Completions ChatGPT-4.1 API along with a structured prompt and the associated abstract text and metadata (one API call/drug) (**Figure 2**). The API returned an evaluation of pediatric safety based on the provided abstracts with temperature set to 0 to enhance reproducibility (**Figure 3A**). All article summaries, including the abstracts that were passed to GPT-4.1, are available on GitHub (**Supplemental Figure 1**).

**Figure 2.**
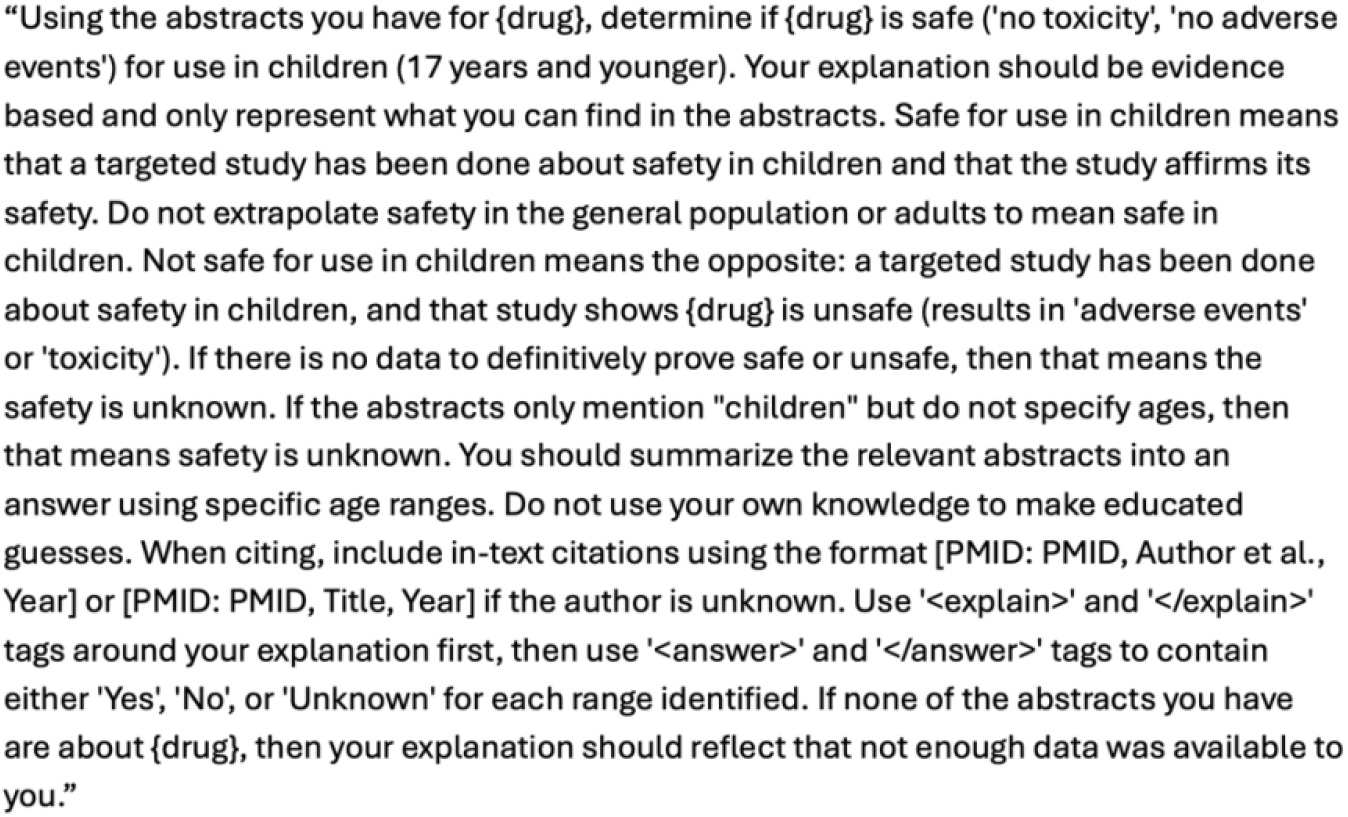
LLM Prompt. GPT-4.1 was passed the top 25 abstracts as determined by cosine similarity and instructed with this prompt, which asked PubChat to make determinations about pediatric safety for each drug based on evidence within the abstracts. Each determination was to be accompanied by a structured explanation and in-text citations, which were matched to metadata in Qdrant.

**Figure 3.**
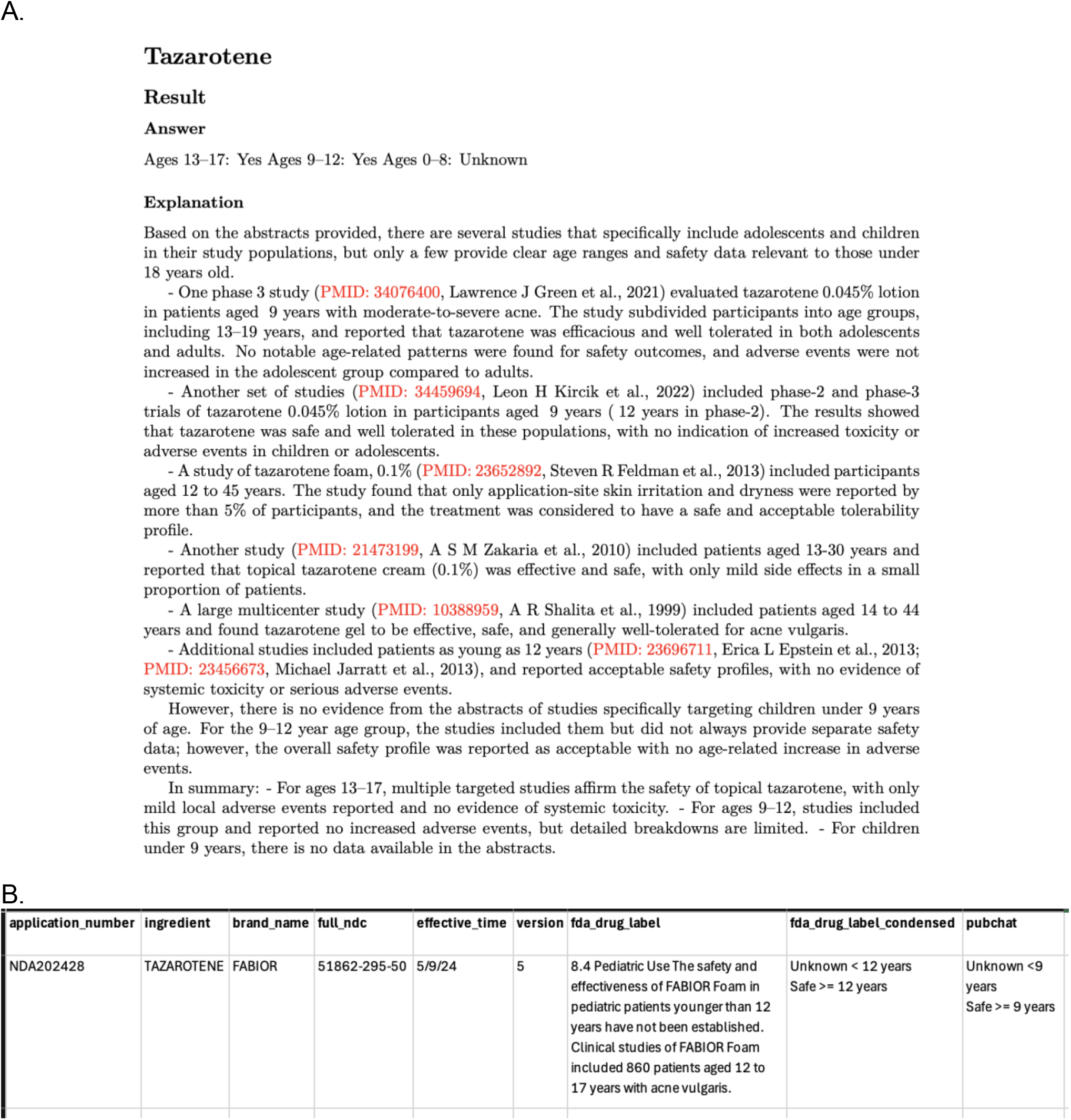
Example Pediatric Safety Evaluation of Tazarotene. Abbreviated Model Output of Tazarotene Pediatric Safety Profile (**3A**). Abbreviated model output of the pediatric safety profile for tazarotene, summarizing safety findings across clinical trial data and literature evidence (**3B**). Comparison of Drug Label to Model Output. Comparison of FDA-approved pediatric labeling with the model-generated output. Drug labeling was retrieved from the FDA Online Label Repository and parsed using an automated pipeline that isolates the Pediatric Use (8.4) section of the Prescribing Information. Age-specific statements, such as “Safety and effectiveness have not been established in patients <12 years,” were manually mapped to categorical labels (“Safe”, “Unsafe”, and “Unknown”. PubChat output was then aligned with the FDA label to highlight areas of concordance (e.g., safety >= 12 years) and divergence (e.g., model identifies safety supported in 9–11 years based on literature but absent from FDA-approved labeling).

### 2.4. Validation

To validate the results, PubChat-generated classifications were compared to the pediatric labeling data information obtained from the FDA database (**Figure 3B**). Labels were manually categorized to fit within constrained classifications, including “Safe”, “Unsafe”, and “Unknown” for each age category. If a drug was indicated for multiple conditions and ages, the drug was labelled as “Safe >= X years old”, where X is the younger of the ages. This categorization was necessary because the content and structure of labeling is not uniform across drugs. These variations arise from several regulatory factors, often between brand and generic formulations, as generic manufacturers are only required to demonstrate bioequivalence rather than replicating the pediatric studies completed by the original brand manufacturer.^14^ Additionally, some labels include specific age range information, for instance, “the safety and efficacy of {drug} has been established for {indication} for children 12 years and older,” while other labels are vaguer. This label would be categorized to “Safe >= 12 years old” and “Unknown < 12 years”.

To establish a baseline to compare the FDA and PubChat labels, ground truth was determined by evaluating the available evidence for each drug and designating the most appropriate label. Each “explanation” and “answer” was manually evaluated to ensure its contents were consistent with the original abstract and each other (**Figure 3B**). Determinations of ground-truth are available in **Supplemental Table 2**. A custom Python script was also used to match in-text citations in the “explanation” with original abstract metadata, and no hallucinations were detected.

### 2.5. Class Imbalance Adjustment

The random sample did not contain any overtly unsafe drugs, which introduced a class imbalance into the evaluation. To address this limitation, we conducted a separate analysis of five drugs that had at least one contraindicated age group in children.

## 3. Results

PubChat was evaluated for 80 FDA-approved drugs with pediatric labeling information. Across the 80 drugs, the FDA labels were categorized as 14 (17.50%) “Safe”, 21 (26.25%) “Unknown < X Years, Safe >= X Years”, 43 (53.75%) as “Unknown”, and 2 (2.5%) as “Other”. PubChat predicted 18 (22.5%) to be “Safe”, 21 (26.25%) “Unknown < X Years, Safe >= X Years”, 6 (7.5%) “Unsafe”, 34 (42.5%) “Unknown”, and 1 (1.25%) “Other” **(Table 1)**.

**Table 1.**
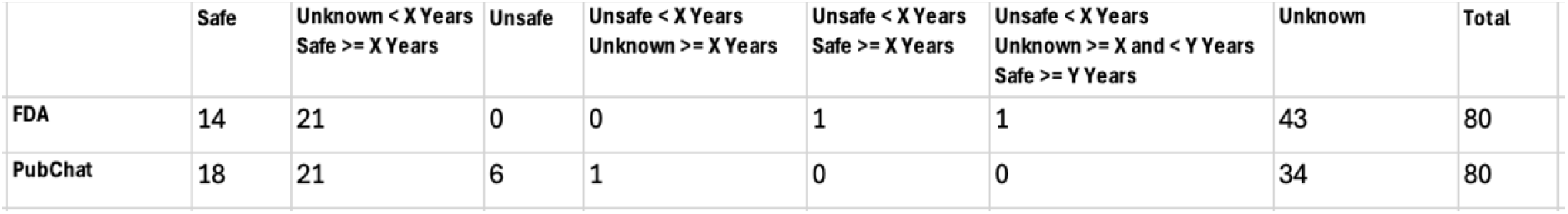
Summary of Pediatric Labeling Results. Each row represents a pediatric labeling outcome category, defined by whether a drug is considered “Safe”, “Unsafe”, or “Unknown Safety” at specific pediatric age thresholds (e.g., “Unknown < X Years, Safe >= X years”). Columns report the count of drugs assigned to each category by FDA labeling versus PubChat.

Concordance was calculated between the two groups and was defined as an exact match between labels. If a drug had more than one category for its label, all were required to match; for example, “Unknown < 12 years, Safe >=12 years” would not match “Unsafe < 12 years, Safe >= 12 years”. Concordance was established in 33 of 80 drugs (41.25%). Of these 33 drugs, 24 (72.7%) were classified as “Unknown” in both groups.

To compare the granularity of safety by age range, age resolution was defined as the ability to provide a definitive safety label beginning at the youngest possible applicable age that matched ground truth. For instance, if the FDA labeled the drug as “Unknown < 12 years, Safe >= 12 years”, and PubChat labeled the drug as “Safe”, and PubChat’s label matched the ground truth, then PubChat was determined to have greater resolution for that drug. Across the 80 sampled drugs, the FDA demonstrated greater resolution in 17 drugs (21.25%), and PubChat demonstrated greater resolution in 15 of 80 (18.75%) of drugs.

PubChat produced overly permissive classifications in 8 of 80 cases (10%) (**Table 2**). In each instance, some supporting evidence of safety was present, but PubChat misinterpreted this information, most often by assuming that the evidence applied from birth. However, in four of these cases, butorphanol tartrate, tranexamic acid, methylprednisolone acetate, and pyridoxine hydrochloride, PubChat did produce compelling safety evidence that was not apparent by reading the drug’s entry in the FDA database, suggesting that the FDA label could be updated to better reflect this evidence. Thus, PubChat identified a total of 19 (23.75%) cases with supporting evidence that the FDA age recommendation was potentially too conservative. However, since PubChat did not correctly summarize the studied ages, these entries were classified as overly permissive.

**Table 2.** Overly Permissive Classifications of Pediatric Labeling Results. PubChat incorrectly classified eight of eighty drugs as safe or conditionally safe. In each case there was supporting safety evidence, sometimes not identified by the FDA, but PubChat mistakenly assumed that the evidence applied from birth or extrapolated a small sample size study to assume safety.

| Active Ingredient | FDA | PubChat | Discrepancy Reason |
| --- | --- | --- | --- |
| ALITRETINOIN | Unknown | Unknown < 6 years<br>Safe >= 6 years | One study does support safety, but sample size is small. |
| AZATHIOPRINE SODIUM | Unknown | Safe | The studies do not come to a clear consensus. |
| BUTORPHANOL TARTRATE | Unknown | Safe | There is supporting evidence, but not from birth. |
| TRANEXAMIC ACID | Unknown | Safe | There is supporting evidence, but not from birth. |
| METHYLPREDNISOLONE ACETATE | Unknown | Safe | There is supporting evidence, but not from birth. |
| PYRIDOXINE HYDROCHLORIDE | Unknown | Safe | There is supporting evidence, but not from birth. |
| VIGABATRIN | Safe >= 1 month<br>Unknown < 1 month | Safe | There is supporting evidence, but not from birth. |
| CEFUROXIME SODIUM | Safe >= 3 months<br>Unknown < 3 months | Safe | There is supporting evidence, but not from birth. One study does mention neonates, but it doesn't specify ages. |

In the sample of 5 drugs with at least one age-related contraindication, PubChat overestimated safety in a single case, Tramadol (**Supplemental Figure 3**). This misclassification arose from reliance on an abstract that did not fully represent its study’s findings, illustrating a potential limitation of relying on abstracts to assess safety.

## 4. Discussion

This study explores the effectiveness of applying a semantic search combined with LLM summarization to the evaluation of pediatric drug safety. The integration of embeddings with vector search enabled scalable retrieval of relevant biomedical literature, while GPT-4.1 provided structured, interpretable evaluations of safety in children. PubChat demonstrates how natural language processing methods can help bridge longstanding gaps in pediatric drug safety, especially in contexts where regulatory labels are not completely current or are lacking specific information.

While PubChat did not outperform the FDA, it did identify 19 drugs where safety information provided by the FDA could be enhanced. Of these, PubChat provided greater age resolution in 15 drugs. In the remaining 4, it found novel research that suggested safety but made an incorrect safety classification. These findings suggest that PubChat is a valuable, complementary tool for clinicians and researchers who rely on updated research to make informed recommendations. An important advantage of

PubChat is its ability to re-rank the most relevant abstracts from a vector database. While its higher-level synthesis of summaries occasionally introduces errors, the tool still provides clinicians and researchers with a valuable starting point for interpreting emerging evidence. Thus, careful human review, especially when model outputs diverge from FDA labeling is essential, particularly for drugs with known adverse effects in children.

This study has several important limitations. First, PubChat relies on information provided in abstracts, which do not always include explicit references to the studied ages. Additionally, abstracts may not fully capture relevant safety information. Also, while PubChat is knowledgeable through 2024, articles published after this date are not included, and future versions should include real-time updates.

Overall, PubChat provides a solution for accessing and summarizing medical literature in the context of the pediatric population. Its primary benefit lies in its ability to assist in evaluating the safety profiles of off-label and investigational drugs which are disproportionately prescribed to children. When used in conjunction with existing FDA labeling, PubChat could serve as a powerful tool in assisting clinicians in making more informed and evidence-based recommendations for pediatric patients, particularly in rare-disease instances where drug repurposing can lead to beneficial treatments.

## Data Availability

Available on github.

https://github.com/avasmith98/Pediatric_Preprint

## 5. Supplementary Materials and Data Availability

All code and supplemental materials are available at: https://github.com/avasmith98/Pediatric_Preprint

## Notes

* This work is supported by the ARPA-H-BDF program.

### Competing Interest Statement

The authors have declared no competing interest.

### Funding Statement

This study was funded partly by the ARPA-H Biomedical Data Fabric program.

